# Health Management Information System Data Quality and its associated factors in Addis Ababa Public Hospitals, Ethiopia, 2022.A cross-sectional study

**DOI:** 10.1101/2025.01.19.25320816

**Authors:** Hiamanot Abebe, Menelik Legesse, Yohannes Godie, Dires Birhanu, Fekadeselassie Belege, Fasil Menbere, Yitayal Guadie

## Abstract

Health management information system (HMIS) is a core health system building block designed to provide important data for continuous quality improvement at all levels of decentralized healthcare administration. Poor data quality at the lower administrative level and health facilities, which are the source of most data used for decision-making in the health sector remains a challenge. This challenge remains high in developing countries, especially in Sub-Saharan Africa.

**Objective:** To assess the level of Health management information system data quality and associated factors among of public hospitals in Addis Ababa, 2022.

**Methodology:** Institutional-based cross-sectional study design was conducted from June to September 2022. Study hospitals were selected using the lottery method and the participants were recruited based on their availability till the required sample was fulfilled variables that have p-value ≤ 0.25 were taken into the multivariable model to control possible confounders. The level of statistical significance was declared at p-value ≤ 0.05. The odds ratio with 95% CI was used to determine the direction and strength of association between the dependent and independent variables.

**Result:** The overall level of HMIS data quality was 78% with completeness at 67%, accuracy at 67%, and timeliness at 100%. Presence of training program (AOR=5.6, CI:2.01, 15.79), notify as job descriptions and report documents clearly (AOR=2.7, CI:1.334, 5.378), formats easily understandable AOR=4.1, CI:1.991,8.256), routine meetings how to improve HMIS data quality(AOR=2.7, CI:1.345,5.417), incentives for HMIS process (AOR=2.3, CI:1.128,4.675) and user-friendly register/report forms (AOR=4.1, CI:1.991,8.256) were the identified factors associated with HMIS data quality.

**Conclusion and recommendation:** The data quality of HMIS was lower than the nationally expected level (85%). Thus, regular and comprehensive training of health care workers, clear job descriptions and HMIS report documents, user-friendly and easily understandable formats, routine meetings to discuss HMIS quality improvement, and incentives for HMIS processes shall be implemented to increase the data quality.

## Introduction

Health information system is among the six building blocks of the health system that play crucial roles in helping the health system. It provides equitable quality health care through the proper use of quality data in service provision, planning, performance monitoring, evaluation, and evidence-based decision-making(Ministry of Health, 2020).

High-quality data on the services provided by health facilities are necessary to make informed decisions regarding resource allocation, planning, and programming (Gebeyehu Bulcha et al, 2019). However, this potentially rich source of data is often overlooked in low- and middle-income countries, because of limited completeness, timeliness, representativeness, and accuracy (Wilson E, et al, 2017).

Studies in Sub-Saharan Africa have reported challenges with data quality, including completeness, timeliness, accuracy, consistency and poor utilization of HMIS tools which are compounded by human, health system, and infrastructure factors. Excessive data demand, large number of reports, frequent changes in HMIS tools, changes in organization structures or human resources, lack of effective systems to monitor quality and absence of standards guidance to measure data quality contribute to poor quality (Rumisha et al, 2020).

According to the assessment conducted on HMIS data quality and information use in Ethiopia, content completeness, reporting timeliness and accuracy were 39%, 73% and 76% respectively (Mastewal Solomon et al, 2021). Poor data quality at the lower administrative level and health facilities, which are the source for the majority of data used for decision-making in the health sector remains challenging (Mastewal Solomon et al, 2021).

Different studies found that Ethiopian HMIS data quality was below the national expectation level (85%) with major data quality dimensions ((Teklegiorgis et al, 2016), (Kebede M et al, 2020), (Shama AT et al, 2021)). This indicates that the need to assess the level of HMIS data quality and its determinant factors so important to improve HMIS data quality and to generate evidence-based decisions for responsible bodies. Hence, the objective of the study was to assess the level of HMIS data quality and its associated factors in Addis Ababa Public Hospitals, Ethiopia. The study hinged on the WHO performance of routine information system management (PRISM) conceptual framework which hypothesized that technical, behavioral, and organizational factors affect HMIS data quality in Addis Ababa public hospitals (USAID, 2009) (Mastewal Solomon et al, 2021).

## Materials and Methods

### Study Design, Area & Period

This facility-based descriptive cross-sectional study design was conducted in Addis Ababa public hospitals from June to September 2022.

Addis Ababa; the capital city of Ethiopia and headquarters of Africa Union is a city comprising 11 sub-cities. In Addis Ababa, there are 12 public hospitals, 97 government health centres 25 private hospitals, and 980 Specialty clinics. Among these, six hospitals (Gandhi Memorial Hospital, Yekatit 12 Hospital Medical College, Zewditu Memorial Hospital, Ras-Desta Damtew Memorial Hospital, Menillik II Referral Hospital, Tirunesh Beijing General Hospital) are governed by the Addis Ababa City Administration Health Bureau and the remaining five (St’ peter Specialized hospital, St Paul’s Hospital millennium medical college hospital, Alert hospital, Amanuel hospital and Eka Kotebe hospital are governed by the federal ministry of health and, one university hospital (Tikur Anbessa Specialized Hospital (Black lion hospital)) is governed by Addis Ababa University(Yasseri T et al, 2012).

By applying random selection using the lottery method, the study was conducted in St. Peter Specialized Hospital (SPSH), Zewditu Memorial Hospital and Menilik II Memorial Hospital (MIIMH).

All health care providers in Addis Ababa public hospitals were a source of population whereas; all health care providers in selected Addis Ababa Public Hospital were the study population. A single study subject /health care provider/ was the study unit/ element. Being a health care provider in selected public hospitals was an inclusion criteria and being sick, on internship, and having work experience less than six months were exclusion criteria.

### Sample size determination

The sample size (n) is determined by using a single population proportion formula and calculated by taking the following statistical assumptions.

p= the proportion taken from the research done at Addis Ababa city administration was (76%)(Haftu B et al,2021).
Z α/2 = the corresponding Z score of 95% CI
d= Margin of error (5%)
N= Sample size

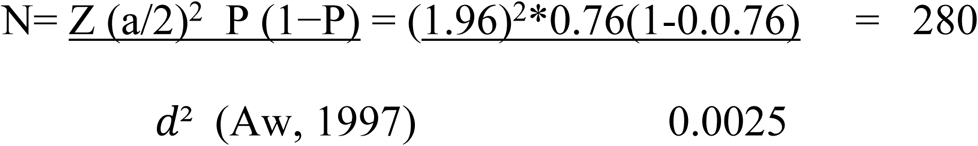

Based on this, from each institution of human resource directorate, the staff distribution in SPSH is 827 health care providers, MIIMH 1000, Zewditu Memorial Hospital 605 were present. This indicates that the source population is less than 10000, correction factors are used to estimate the final sample size required.

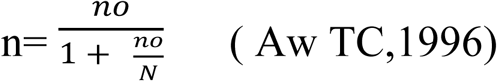

nf =final sample size
N=total number of source population is 2432

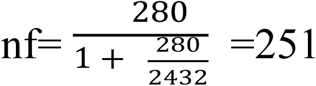

Taking the non-response rate, the total sample size for this study is 251+25=<u>276</u>

Second objective sample size calculation for different factors associated with HMIS data quality:

For the second specific objective sample size is calculated considering two population proportions using Epi-info version 7.0. Factors that were significantly related to the dependent variable were used based on the following assumptions 95 % level of confidence, 80% power, and finally the two-sample size was compared and a formula that provides maximum sample size was used (Table-1).

**Table 1:** Calculated sample size for factors associated with HMIS data quality among Addis health care providers.

| **Factors** | **CI** | **Power (1-β)** | **Ratio** | **Proportion of outcome** | **OR** | **Sample size(n)** | **Final sample size (by adding 10% non-responses...)** | **Source** |
| --- | --- | --- | --- | --- | --- | --- | --- | --- |
| **Training** | **95%** | **80%** | **1** | **49** | **2.9** | **138** | **152** | **(Shama et al., 2021)** |
| **Feedback from higher report receivers** | **95%** | **80%** | **1** | **48.7** | **3.1** | **124** | **136** | **(Shama et al., 2021)** |
| **seek feedback** | **95%** | **80%** | **1** | **75.3** | **2.5** | **300** | **330** | **(Teklegiorgis et al,2016)** |

### Sampling technique

From the twelve hospitals, three hospitals, Zewditu Memorial Hospital, St. Peter Specialized Hospital, and Menillik II Memorial Hospital were selected using the lottery method. A list of healthcare providers was obtained from the service units of each hospital. Based on this, from each institution’s human resource directorate, the staff distribution in SPSH was 827(333-nurse, 201-general practitioners, 188-midwifes and 105 -seniors) MIIMH, 1000(415-nurse, 295-general practitioners, 168-midwifes and 122-seniors) ZMH 605(258-nurse, 193-general practitioners, 76-midwifes and 78-seniors). Each study participant was selected randomly till the desired number of participants from health professionals that are involving in clinical activities in each hospital was achieved (based on availability) **(Table 2).**

**Table 2:** Sample size allocation per hospital and professional distributions.

| Selected<br>Hospitals | Total population |  |  | Sampled |  |  |  | Sub-<br>total |
| --- | --- | --- | --- | --- | --- | --- | --- | --- |
|  | Total<br>Specialist | GP | Midwives<br>And<br>Nurses | Total<br>Specialist | GP | Midwives<br>and nurses | Total |  |
| **MIIMH** | **122** | **295** | **583** | **17** | **40** | **79** | **136** | **330** |
| **ZMH** | **78** | **193** | **334** | **11** | **26** | **45** | **82** |  |
| **SPSH** | **105** | **201** | **521** | **14** | **27** | **71** | **112** |  |

### Study variables

#### Dependent variable (Out-come variable)

HMIS data quality

#### Independent variable

Socio-demographic variables: Age, sex, position and work experience in current position.

##### Technical/structural factors

The complexity of the reporting, Formats and procedures, Individual/Behavioural factors: Motivation to do data quality, perception towards HMIS Data quality and data quality checking skill.

##### Organizational factors

Governance, Supervision, training provided to staff and culture of information

### Questionnaire Development and validation

Self-administered structural questionnaires and record reviews were adapted from the PRISM assessment tools version 3.1 and current Ethiopian HMIS user guidelines (USAID, 2009). The tool was externally validated by researchers and health service quality experts.

### Data collection, procedures, and Quality

The tool was prepared to fit with the local context and it mainly contains health facility tools for routine data quality to assess the accuracy, completeness, and timeliness of HMIS data. The data was collected by self-administered questionnaires and record reviewing by using HMIS tools adopted from PRISM assessment tools version 3.1(USAID, 2009). The tool was prepared in English and fitted with the study’s objectives, which contains health facility tools that assess the accuracy, completeness, and timeliness of HMIS data. The self-administered structured questionnaire was divided into different parts socio-demographic factors, organizational factors, behavioral factors, and technical factors. The data collection procedure was done after informed written consent had been taken.

To ensure the quality of the data, Six (6) professionals with a Master of Science (MSc) were recruited and two days of training were given to two supervisors and four data collectors. The training was on the aim of the study, procedures, data collection techniques, data contamination, research bias, research ethics, consent, and rights of study subject confidentiality. Pre-tested on 5% (n=33) was held in Yekatit 12 Medical College Hospital. After data collection, each questionnaire was coded and checked for completeness and consistency prior to data entry. The data were checked for missed values, inconsistencies and outliers.

### Data analysis procedure

Data were entered using Epi-data version 7.1 and exported to SPSS Software version 26 for analysis. Bi-variable logistic regression analysis was used to see the association between each independent variable & the outcome variable. Multiple logistic regression analysis was carried out to determine the effect of various factors on the outcome variable. All variables with a p-value ≤ 0.25 in the bi-variable logistic regression analysis were included in to the multi-variable model to control for potential confounding factors. The degree of association between dependent and independent variables was assessed using odds ratio with a 95% confidence interval and p-value ≤0.05. Finally, the result was presented by texts, tables, and figures.

## Result

A total of 330 questionnaires were distributed to healthcare providers working at three public hospitals in Addis Ababa, and 324 questionnaires were returned, resulting in a response rate of 98.2%. Among the respondents, 215 (66.4%) were female, and more than half of the participants, 236 (72.8%), were nurses, followed by 50 (15.4%) midwives and 28 (8.6%) doctors. Ninety respondents (27.7%) reported receiving any training program about the HMIS. The majority of respondents, 273 (84.3%), were staff members, and 29 (8.9%) were case managers. Nearly half of the participants (48.5%) earned less than 7,500.00 ETB, while 167 (51.5%) earned between 7,500.00 and 15,000.00 ETB, and none (0%) of the participants earned a monthly salary above 15,000.00 ETB **(Table 3).**

Regarding participant’s Work experience

Approximately 196 (60.49%) of the respondents had less than 5 years of work experience, followed by 96 (29.63%) with 5 to 9 years of service, and 32 (9.88%) with more than 10 years of experience (Figure 1).

**Figure 1:**
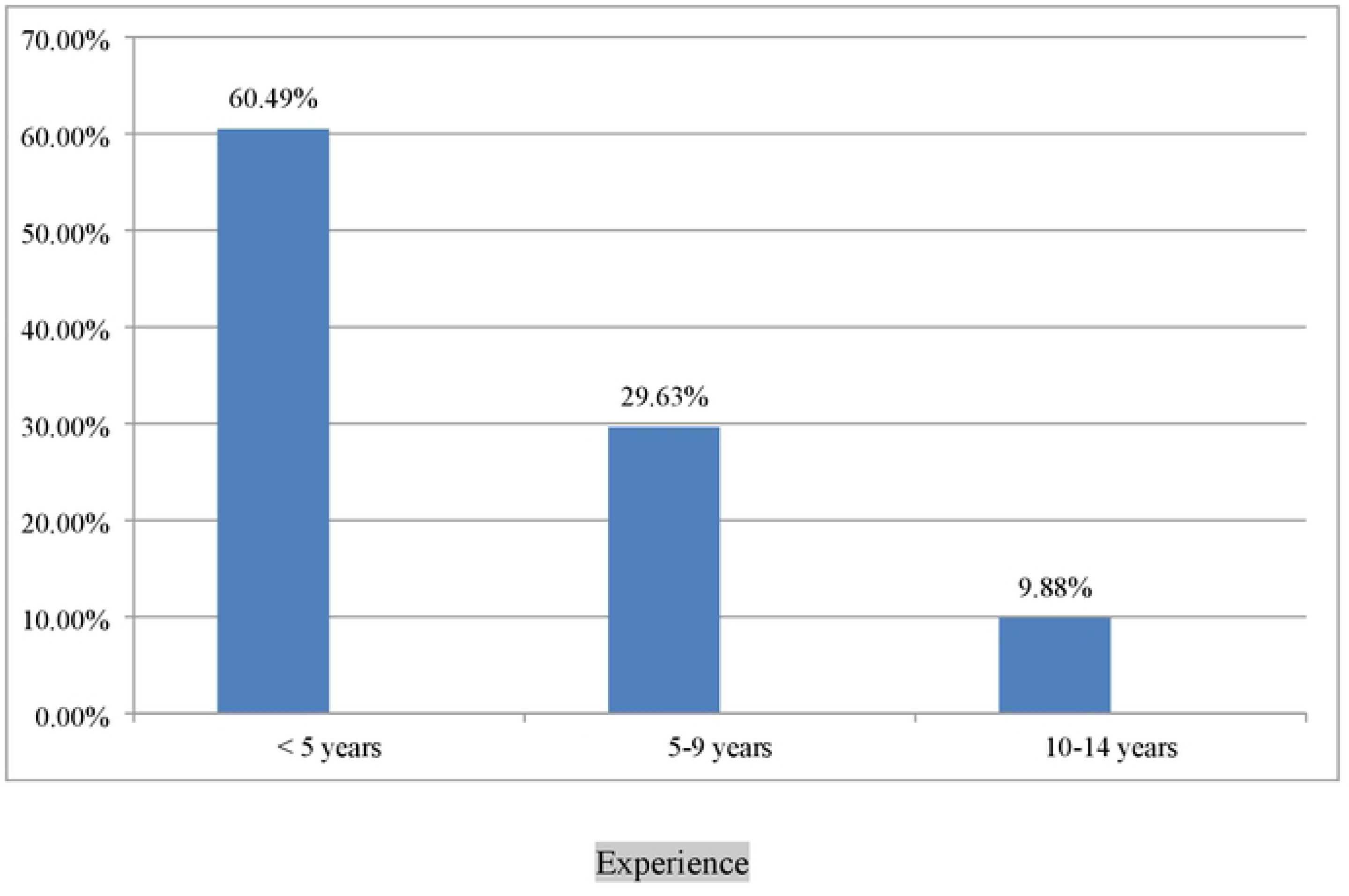
Work experience of health care worker public hospital Addis Ababa City, 2022

### Technical factors

From the total respondents, 170 (52.5%) reported that their facility had data management and quality guidelines, as well as a standard set of HMIS indicators, including case definitions, in their working area. Among the respondents, 214 (66%) stated that they register all eligible activities daily in the HMIS register, and 243 (75%) had received job descriptions that outline their responsibilities for recording, reporting, and compiling HMIS data(**Table 4).**

### Organizational factors

Out of 324 respondents, 208 (64.2%) had participated in the aggregation or compilation of HMIS data from the data sources, and 216 (66.7%) of them agreed that the registration and reporting formats were easily understandable. The majority of respondents, 289 (89.2%), reported that their departments had officially assigned an HMIS focal person responsible for compiling, organizing, and reviewing HMIS data before submission to a higher level. Regarding incentives, 121 (37.3%) of the respondents agreed that incentives could motivate and create a sense of ownership regarding HMIS activities. Additionally, 289 (89.2%) of the total respondents reported that they had an HMIS focal person responsible for reviewing data before submission to the next level **(Table 5).**

### Individual factors

Individual and behavioral factors were assessed through the respondents’ perceptions, attitudes (motivation) towards HMIS use, skills regarding HMIS, confidence levels to perform HMIS tasks, and related activities for HMIS data. Out of the total respondents, 302 (93.2%) had the knowledge to correctly calculate percentages and rates from HMIS tools, and 263 (81.2%) could explain the findings of HMIS reports and their implications. Approximately 250 (77.2%) respondents reported that they could collect reliable data from different HMIS data sources, and 245 (75.6%) of respondents agreed that they had used the generated HMIS data for day-to-day activity evaluation. (Table 6).

### Level of data quality from accuracy perspectives

Out of the 12 public hospitals in Addis Ababa, three hospitals were studied for data quality, evaluating the dimensions of accuracy, completeness, and timeliness. Five data elements or indicators were assessed for data accuracy. Disease reports and HMIS registration books from June 2022 and August 2022 were checked to enable the assessment of the most recent data quality status. The data elements evaluated were newly diagnosed diabetes mellitus, severe acute malnutrition, all types of pneumonia, confirmed tuberculosis cases, and the total number of births attended by health workers. From the three selected public hospitals, 2 (67%) reported within the acceptable range (90-110%), but 1 (33%) of the public hospitals had under-reports (<90%), and no public hospitals had over-reports. The overall accuracy of the data was 67 %(**Table 7).**

### Level of data quality from completeness perspectives

Content completeness was assessed by checking the service delivery reports from two months to determine whether the required data elements in the report form were filled or the data were complete. Based on this assessment, the overall content completeness was recorded as 67 % **(Table 7).**

### Level of data quality from timeliness perspectives

The timeliness of the HMIS reports was assessed by checking whether the HMIS data reporting by the public hospitals met the predetermined national deadline for the reporting period to submit reports to the next level. From the three selected public hospitals, all (100%) had records and reports containing the date of submission to the next level, indicating that the HMIS reports were sent within the acceptable time period. Therefore, the overall HMIS data timeliness was 100 **% (Table 7).**

### Total quality of HMIS data in three dimensions

Based on the assessment of the three dimensions of HMIS data quality - timeliness, completeness, and accuracy - the overall data quality (mean score) of the public hospitals was 78%. Of the three study facilities, Zewiditu Memorial Hospital has the lowest data accuracy, while St. Peter’s facility has the lowest data completeness **(Figure 2).**

**Figure 2:**
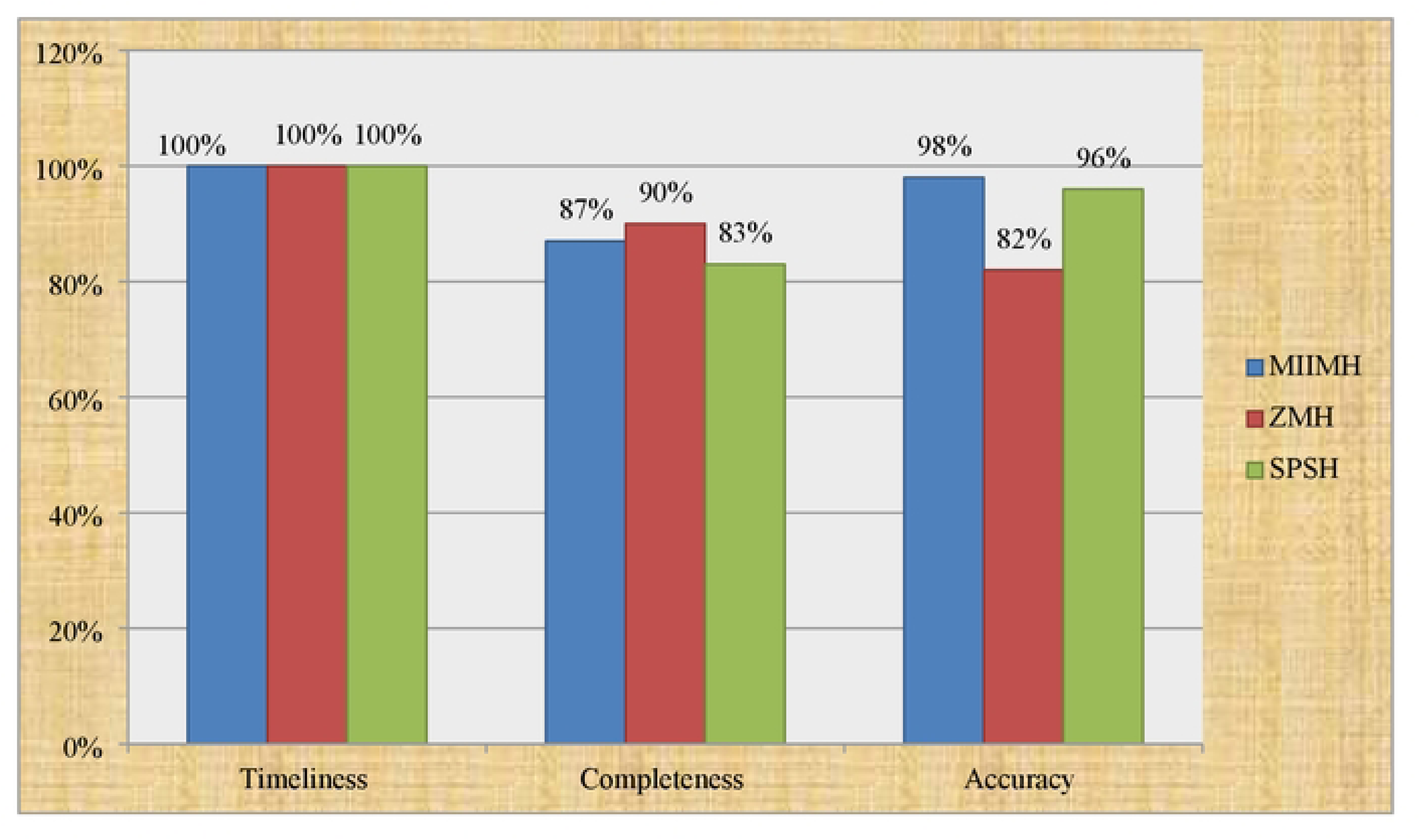
Overall HMJS data quality for the study conducted to assess HMJS data quality and its associated factors in public hospitals of Addis Ababa, Ethiopia, 2022.

### Bi-variable and multi-variable analysis

All variables were first tested using bi-variable logistic regression to select candidate factors for the multi-variable logistic regression analysis. Variables with a p-value less than 0.25 in the bi-variable logistic regressions were then exported to the multivariable logistic regression model to control for possible confounders. With multiple logistic regressions result, training participation (AOR: 5.6, CI: 2.01, 15.79), job descriptions deal with the responsibility for recording and compiling HMIS data with (AOR: 2.7, CI: 1.334, 5.378), availability of user-friendly HMIS registrations and report formats (AOR: 4.1, CI:1.991,8.256), any incentives for HMIS process (AOR: 2.3, CI 1.128,4.675), routine meetings for reviewing HMIS data quality (AOR: 2.7,CI:1.345,5.417) and practice of using generated HMIS data for days to-day activities evaluation (AOR: 2.8, CI:219,6.380) were the identified factors associated with HMIS data quality **(Table 8).**

## Discussion

In this study, the overall data quality was 78%, which is higher than the findings reported from previous studies conducted in Eastern Ethiopia (75.3%), West Gojam (74%), and Harari regional state (51.35%) (Shama et al., 2021). The possible explanation for this difference might be due to variations in the professional distribution, as this study included all the professions working in the departments, as well as the availability of basic instruments and human resources to carry out the data quality assessment. It’s also possible that the study setting has implemented more robust HMIS data quality improvement strategies compared to the areas covered in the previous studies. On the other hand, the result of this study was lower than the study conducted in the Hadiya zone of Southern Ethiopia, which reported an overall data quality of 82.5% (Mastewal Solomon et al., 2021). The possible scientific explanation for the difference might be due to the fact that number of cases can matter to have time for the professionals to fill HMIS register and compile all report neatly. This is generalized by the fact that the more cases flow, the shorter the time for the professionals to organize basic data quality activities. The Hadiya zone may have had more favourable contextual factors that enabled higher data quality compared to the setting of the current study. This could include factors like stronger health system infrastructure, more robust HMIS implementation, or higher levels of staff capacity and motivation.

In this study the accuracy of data was found to 67% which was higher than studies conducted in East Wollega and Harargi(Shama et al,2021). The possible explanation for this difference might be due to professional distribution difference in which this study includes all the professions working in public hospitals, availability resources which enhance for data quality. The study setting may have cultivated a more positive organizational culture around HMIS data use and quality could be another justifications.

This finding was nearly similar with the study conducted in Southern Ethiopia, Hadiya zone(76%) (Mastewal Solomon, 2018). The possible scientific explanation for this finding might be due to the similar type of facilities and use indicators to assess data quality. Ethiopia has national HMIS standards and guidelines that are intended to be applied consistently across the country. If both of these study settings have effectively implemented these common HMIS policies and practices, it could contribute to the convergence in data accuracy levels.

Regarding completeness, the magnitude was found to be 67% which was lower than national expectation (85%) and a study conducted in Southern Ethiopia, Hadiya zone 83.3%(Mastewal Solomon, 2018). The possible scientific explanation for this finding might be due to the fact that number of cases can matter to have time for the professionals to fill all HMIS data components. There may be specific gaps or weaknesses in the local HMIS data management practices, such as inadequate data collection, entry, verification, or reporting procedures. But greater than a study conducted in Harari region (60%)((Shama et al, 2021)). Possible reasons may be due to increased knowledge of respondents about the implications of an incomplete data on a report format since information revolution national agenda was launched nationally followed by different trainings related data quality.

In this study timeliness of the HMIS data was 100% which is greater than the finding extracted from a study conducted in Hadiya zone (73%) and Harari (93.7%)((Shama et al,2021)). The possible scientific explanation for this difference might be due to the fact that using soft copy format to send all HMIS report online and close follow up of deadline of report by department head, assigned HMIS focal person free from other hospital tasks at public hospital level, continuous accessibility of resource (internet, transport) and it may also due to increase knowledge of respondents about the exact date nationally reported time period and deadline.

Health care providers who participated in HMIS training programs were 5.6 times more likely to organize high-quality HMIS data compared to their counterparts (AOR = 5.6, CI: 2.012, 15.787). This finding is similar with the study done in Hadiya zone and Harari region(Shama *et al*, 2021). This might be due to training for health care providers create the chance being user friendly with different registers, tally sheets and report formats and also increase their awareness about the importance of quality HMIS data. HMIS training programs equip health care providers with the necessary knowledge, skills, and competencies to properly collect, record, and organize health data. This enhanced capacity directly enables them to produce higher-quality HMIS data. Training programs often emphasize the value of using HMIS data for planning, monitoring, and evaluation. When health care providers understand the practical applications of data, they are more likely to invest effort in ensuring its quality.

Health care providers those who have received job descriptions deals about the responsibility for recording, organizing, aggregating and reporting of HMIS data were 2.7 times more likely to have a good quality of HMIS data as compared to health care providers those who did not know their duties, roles and responsibilities towards HMIS data (AOR=2.7,CI: 1.334, 5.378). This finding was in line with the study conducted in West Gojam(Chekol A et al,2021) and Addis Ababa(sebsbie A,2020).This might be due to developing sense of responsibility and well understanding their scope of practice. When HMIS responsibilities are explicitly included in job descriptions, it demonstrates the strategic importance of high-quality data for the organization’s objectives which signifies an organizational commitment to HMIS and data quality. This support, along with any associated resources and supervision, enables health care providers to fulfil their data management duties more effectively.

Health workers that agreed complexity of registrations, tally sheets and report formats were 4.1 times more likely to have good quality HMIS data compared to those health workers who did not agree on user friendly registrations, tally sheets and report formats (AOR 4.1, CI:1.991, 8.256). This finding was comparable with studies conducted in Jimma (Tilahun B et al,2021) and West Gojam(Chekol A et al, 2021).This might be because of low awareness and practice towards registrations, tally sheets and report formats the reasons related to non-understand ability, ambiguity.

In this study, provision of any incentives for HMIS process/ Staff get rewarded for good work on data management had a positive association with good HMIS data quality in which health workers who get rewarded for good work on data management were 2.3 more likely to practice good quality of HMIS data formats AOR 2.3,CI:1.128, 4.675). This finding was comparable with studies conducted in East wollega (Kebede M et al,2020). This might be since health professionals who get rewarded for good work on HMIS data management had the commitment to fill and understand all necessary tally sheets, registration and report formats develop skill to compile analyses data, and utilize quality of generated quality HMIS data in the routine day-to-day activities. Providing incentives or rewards for good HMIS data management work can significantly boost the motivation and engagement of health workers. When they feel their efforts are recognized and valued, they are more likely to prioritize data quality. The introduction of rewards can also create a sense of healthy competition among health workers, encouraging them to learn from each other’s best practices and strive to improve their individual and collective HMIS data quality

This study also noted that health care provider who used generated HMIS data for their health related to-day activities evaluation is strongly associated with quality of HMIS data. That is health care provider who used generated HMIS data for their health related to-day activities evaluation 2.8 times more likely to compile quality of HMIS data than those health care providers who did not used generated HMIS data for their health related to-day activities evaluation (AOR 2.8, CI:1.219, 6.380). This result is supported by a study conducted in East wollega (Kebede M et al,2020). This might be due to that fact that, when health care providers actively utilize the HMIS data in their practical work and decision-making, it reinforces the value of high-quality data, fostering a data-driven culture and a sense of ownership and accountability for the information they provide. The enhanced understanding of data utility and its role in decision-making further motivates providers to prioritize data quality improvements, while regularly using HMIS data helps cultivate a data-driven mind-set that leads to continuous quality improvement efforts.

This study also revealed that hospitals which had the habit/presence of routine meetings for reviewing HMIS data quality and discuss openly to resolve problems related to HMIS data quality increase 2.7 times fold AOR (2.7, CI: 1.345, 5.417). This result is in line with a study conducted in East Wollega Zone (Kebede M et al,2020).The possible explanation for this finding might be due to the fact that habit of meeting regarding HIMIS can create knowledge and even may be considered as a mandatory rule which increase professional essence. It could also be due to the fact that regular data review meetings foster a culture of data-driven decision-making, where healthcare providers actively engage with the HMIS data, identify areas for improvement, and collaboratively develop strategies to enhance data quality. This collaborative approach not only improves data accuracy and completeness but also promotes ownership and accountability among staff, as they work together to address data-related challenges. Furthermore, the open discussion and problem-solving during these meetings help to build trust, transparency, and a shared understanding of the importance of HMIS data quality, which are crucial for driving long-term improvements within the healthcare system.

## Limitations of a study

Content completeness was assessed only for reporting formats, so it couldn’t represent the completeness of registration and tally sheets. Utilization of self-administered questionnaires may affect the validity of the responses. Using secondary data may underestimate the findings.

## Conclusion and recommendation

Health management information system data quality was lower than the nationally expected level. The overall level of health management information system data quality was 78% with completeness 67%, accuracy 67% and timeliness at 100%. The presence of training program, notify as job descriptions and report documents clearly, formats easily understandable, routine meetings how to improve HMIS data quality, incentives for HMIS process and user-friendly register/report forms were the identified factors associated with HMIS data quality. Hospitals shall strengthen regular and comprehensive HMIS training programs for all healthcare providers, create specific and well defined job descriptions and; conduct routine meetings focused on HMIS data quality improvement that involve all relevant stakeholders. Ethiopian ministry of health shall develop and implement user-friendly and easily understandable reporting formats. Since data is the back-bone of a health care system, non-governmental organizations together with ministry of health shall introduce and implement incentive schemes to motivate healthcare providers by prioritizing data quality.

## Data Availability

all data fully available

## Declaration

## Abbreviation and acronyms

FMOH: Federal Ministry of Health
HMIS: 
HSTP: Health Sector Transformation Plan
KPI: Key Performance Indicators
MIIMH: Menelik II Memorial Hospital
MOH: Ministry of Health
SPSH: St. Peter Specialized Hospital
WHO: World Health Organization
ZMH: Zewditu Memorial Hospital

## Operational definition

Good Quality of data- The data that fits the criteria for the three quality dimensions – accuracy ≥80%, completeness ≥85%, and timeliness ≥85 %

Poor Quality of data- The data that do not fit the three criteria (accuracy <80%, or completeness <85%, or timeliness <85%)

The accuracy of data- was measured by calculating the number from the source document over the number from a report submitted to the next level. Based on 10% tolerance for data accuracy was classified as follows;

✓ verreporting (<0.90 or 90%)
✓ limit (0.90-1.10 or 90%-110%)
✓ (>1.10or110%).

The HMIS data of the facility is considered as accurate if the average was ≥80%

Completeness of data- was the average of the source document or registration content completeness and report content. The data is complete if the average is≥85%

Data is the raw material in the form of numbers or characters.

HMIS- An information system specially designed to assist in the management and planning of health programmer as opposed to improve quality of care.

Timely data- was assessed as a report submission within the accepted period by observing the reporting date on the reporting form of two randomly selected monthly reports. The HMIS data of the facility is timely if the average is≥85%

## Ethical Consideration

Ethical clearance was brought from Menelik II Medical and Health Science College Institutional Review Board with a reference letter of MIIMHS/5/38/12/304 and written permission was obtained from St. Peter specialized hospital. Informed consent was obtained from the participants, after providing information about the purpose, procedures, benefits, risks and confidentiality of the study. The rights of study participants to refuse to answer any questions or stop the interview, or skip any question that they did not want to respond at any time was also respected. All the information given by the respondents was for research purposes only, confidentiality and privacy were maintained by omitting the names of the respondents during the data collection procedure and after data collection information from the study without the participants’ names.

## Consent for publication

All authors approved that this manuscript is eligible for publication and the indexed mothers were well informed as the paper can be published while informed consent was signed.

## Availability of data and materials

The data and other documents used in this study are available from the corresponding author.

## Author’s contribution

**HAL&ML: conceptualization, methodology, data entry, data cleaning, data analysis, writing original paper, DBM and YG,YG,FM and FSB, validation, tool evaluation, methodology, language and statistical reviewing and editing.**

## Acknowledgement

We have special thanks for study subjects, institutions, data collectors, leaders and staff of each study area.

